# Decreased incidence, virus transmission capacity, and severity of COVID-19 at altitude on the American continent

**DOI:** 10.1101/2020.07.22.20160168

**Authors:** Christian Arias-Reyes, Favio Carvajal-Rodriguez, Liliana Poma-Machicao, Fernanda Aliaga-Raudan, Danuzia A. Marques, Natalia Zubieta DeUrioste, Roberto Alfonso Accinelli, Edith M. Schneider-Gasser, Gustavo Zubieta-Calleja, Mathias Dutschmann, Jorge Soliz

## Abstract

The coronavirus disease 2019 (COVID-19) outbreak in North, Central, and South America has become the epicenter of the current pandemic. We have suggested previously that the infection rate of this virus might be lower in people living at high altitude (over 2,500 m) compared to that in the lowlands. Based on data from official sources, we performed a new epidemiological analysis of the development of the pandemic in 23 countries on the American continent as of May 23, 2020. Our results confirm our previous finding, further showing that the incidence of COVID-19 on the American continent decreases significantly starting at 1,000 m above sea level (masl). Moreover, epidemiological modeling indicates that the virus transmission rate capacity is lower in the highlands (>1,000 masl) than in the lowlands (<1,000 masl). Finally, evaluating the differences in the recovery percentage of patients, the death-to-case ratio, and the theoretical fraction of undiagnosed cases, we found that the severity of COVID-19 is also decreased above 1,000 m. We conclude that the impact of the COVID-19 decreases significantly with altitude.

**Highlights:**

1. There is a negative correlation between altitude and COVID-19 incidence on the American Continent starting from 1,000 m above sea level.
2. The transmission rate of SARS-CoV-2 is lower in the highlands than in the lowlands.
3. The severity of COVID-19 decreases significantly with increased altitude.

## 1. Introduction

On March 11, 2020, coronavirus disease 2019 (COVID-19) was declared a pandemic by the World Health Organization (Joshua Berlinger, 2020; World Health Organization, 2020b). In late April, the health crisis began to ease in Asia and Europe (France 24, 2020; Langton, 2020; NPR, 2020), whereas case numbers began to rise in American countries. The first cases of COVID-19 on the American continent were reported in Canada on January 15^th^ and in the United States on January 20^th^. Before February 25^th^, Brazil was the first affected country in Latin America. From that date until July 7^th^, the Pan-American Health Organization (PAHO) (Pan-American Health Organization, 2020) reported 6,004,685 confirmed cases and 268,828 deaths from COVID-19 in the American continent (Pan-American Health Organization, 2020). The spread of the virus was so fast in the region, that it has now become the epicenter of the disease, with United States (1^st^), Brazil (2^nd^), Peru (5^th^), Chile (6^th^), Mexico (9^th^), Colombia (19^th^) and Canada (20^th^) among the top twenty countries with the highest number of confirmed cases in the world on July 7^th^ (Worldometer, 2020b).

Although American countries had more time and information to prepare for the pandemic than did Europe and Asia, with few exceptions, weaker public health systems, late political responses, and complex cultural and social conditions (i.e., poorly respected quarantines in most countries) have led to a major public health crisis on the continent. A crucial aspect for the spread of the virus is overcrowding in large cities. Clear examples of this fact are the cities of New York (8.1 million people - 10,194 people/km^2^ - 214,371 cases) (N. Y. C. Health Department, 2020), Montreal (1,8 million people - 4,517 people/km^2^ - 27,438 cases) (Montréal, 2020), and Rio de Janeiro (6, 7 million people - 5,597 people/km^2^ - 33,695 cases) (Ministerio de Saude de Brasil, 2020) (data as of July 7). Furthermore, this scenario is particularly critical in the poorer districts of those cities, where large numbers of people occupy the same housing units (Rede Brasil Atual, 2020). Remarkably, however, other densely populated metropolises with large slum areas, but located above 1,000 meters above sea level (masl), such as México city (8 million people - 26,000 people/km^2^ - 53,423 cases) (Gobierno de la Ciudad de México, 2020), Bogotá (7,4 million people - 4,310 people/km^2^ - 36,554 cases)(Alcaldía de Bogotá, 2020) and La Paz (2,3 million people - 2,676 people/km^2^ - 4,413 cases)(SEDES La Paz, 2020) (data as of July 7), seem to show lower incidences of COVID-19. This observation is of crucial importance since in American countries, more than 120 million people live at an altitude higher than 1,500 masl (defined as moderate altitude - MA), and more than 35 million people live at an altitude higher than 2,500 masl (defined as high altitude) (Hornbein & Schoene, 2001).

The impact of altitude on a potentially decreased virulence of SARS-CoV-2 was previously reported by our team. In fact, the global data analysis, which included detailed information from the Tibetan Autonomous Region of China, Bolivia, and Ecuador, suggested that the infection rates for the SARS-CoV-2 virus decrease significantly from 2,500 masl (Arias-Reyes, et al 2020). Subsequent reports from other research groups supported this observation (Huamaní, Velásquez, Montes, & Miranda-Solis, 2020; Ortiz-Prado et al., 2020; Rivero & Montoya, 2020). However, being aware that the course of the pandemic changes from day to day and that more detailed statistical analyzes are required, in this new study, we analyzed the epidemiological data from 23 countries in the American continent as of the 23^rd^ of May. Our results show that the incidence of COVID-19, the virus transmission rate, and the severity of COVID-19 decrease significantly starting at 1,000 masl.

## 2. Methods

### 2.1. Data sources

Supplementary Table S1 shows the list of data sources used to collect information from COVID-19 cases through May 23^rd^, 2020. For Bolivia, Brazil, Canada, Colombia, French Guyana, Panama, and USA, we used information about the number of cases per city/county. For Argentina, Belize, Chile, Costa Rica, Cuba, Ecuador, El Salvador, Haiti, Honduras, Mexico, Paraguay, Peru, Puerto Rico, Dominican Republic, Uruguay, and Venezuela, we used information about the number of cases per state/province/departamento.

All locations with reported positive COVID-19 cases were associated with their respective geographic coordinates (latitude and longitude) using the OpenCage Geocoding API (GmbH, 2020). The altitude information for each location was extracted from the WORLDCLIM digital elevation model (Fick & Hijmans, 2017), and the population density data were assigned from the dataset of the CIESIN (Center for International Earth Science Information Network - CIESIN - Columbia University, 2018). In cases where the value of population density was zero, the information was retrieved from the national statistical institute of the corresponding country. The full list of these locations can be found at: https://doi.org/10.6084/m9.figshare.12685610.v1.

### 2.2. Assessment of incidence versus altitude

The number of COVID-19 cases by location was normalized by population density (inhabitants per square kilometer). These data were then organized by intervals of 100 meters of altitude. Finally, the natural logarithm (ln) of each normalized value was calculated. The correlation between the number of COVID-19 cases normalized and logarithmized (ln [cases/population density]) and the altitude was analyzed using a Pearson correlation analysis (n= 51). The difference in COVID-19 incidence below and above 1,000 masl at the continental level was calculated using a bidirectional random block ANOVA. The analyzed variable was the number of COVID-19 cases normalized and logarithmized (ln [cases/population density]); the grouping variable was the altitude (> 1,000 masl or < 1,000 masl) and the block variable was the country (n_>1,000_ masl= 827; n_<1,000_= 3,659). An identical correlation analysis was performed for each of the 23 countries of the American continent (n_Argentina_= 12; n_Belize_= 3; n_Bolivia_= 27; n_Brazil_= 17; n_Canada_= 13; n_Chile_= 14; n_Colombia_= 36; n_Costa Rica_= 20; n_Cuba_= 4; n_Dominican Republic_= 9; n_Ecuador_= 19; n_El Salvador_= 8; n_French Guyana_= 1; n_Haiti_= 8; n_Honduras_= 12; n_Mexico_= 15; n_Panama_= 13; n_Paraguay_= 5; n_Peru_= 17; n_Puerto Rico_= 8; n_Uruguay_= 2; n_USA_= 34; n_Venezuela_= 12).

### 2.3. Evaluation of the virus transmission rate

The evaluation of the SARS-CoV-2 virus transmission rate was performed only for Argentina, Bolivia, Colombia, Ecuador, and Peru, as these countries applied similar strong early quarantines and provided daily epidemiological data at state/province/departamento level.

The COVID-19 data retrieved daily by state/province/departmento in each country was classified into two groups: highlands (>1,000 masl) and lowlands (<1,000 masl). The SEIR model (Susceptible - Exposed - Infectious - Removed) was used to calculate the estimated number of susceptible, exposed, infected and removed (recovered + deceased) individuals (Abadie, Bertolotti, & Arnab, 2020). The number of susceptible individuals was calculated as the total population minus the number of infected and exposed subjects. The initial number of infected was set to 1, and the number of exposed subjects was arbitrarily set to 30 (according to (Birbuet & López, 2020). The contact rate (β), recovery rate (γ), and the rate at which exposed individuals become infected (ε) were calculated as conducted elsewhere (Birbuet & López, 2020). The contact rate was estimated as the product of the “interaction frequency” and the “probability of transmission of the disease” (transmission rate) (Birbuet & López, 2020). Finally, the epidemiological behavior of the disease was modeled, calculating the best transmission rate for highlands (>1,000 m) and for lowlands (<1,000 m).

### 2.4. Assessment of COVID-19 severity

The severity of the disease was estimated based on the percentage of recovered patients and the death-to-case ratio. Indeed, a lower mortality rate per case and a higher percentage of recovered patients suggest a lower severity of the disease (and the opposite). Accordingly, the severity of the disease was estimated for Argentina, Bolivia, Colombia, Ecuador, and Peru, as these countries applied similar containment measures and provided daily epidemiological data at the state/province/departamento level.

The death-to-case ratio and the percentage of recovered patients ([recovered patients/reported cases] * 100) for each country (except Ecuador) were calculated using the data from the last 10 days evaluated (from May 13^th^ to 23^rd^). The underlying rationale is that these parameters stabilize as the pandemic progresses. As this information was not available for Ecuador on the mentioned dates, these values were calculated only from the data of April 29 (the latest data available for this country).

The Wilcoxon signed-rank test was used to determine if the percentage of recovered patients (n_pairs_= 5) and the death-to-case ratio (n_pairs_= 5) were different between the highlands and the lowlands.

### 2.5. Assessment of undiagnosed cases

COVID-19 patients can be asymptomatic or symptomatic. According to the severity of the disease, symptomatic patients are classified as follows: 1) Mild: patients with very mild symptoms and without evidence of viral pneumonia or hypoxia; 2) Moderate: patients with signs of pneumonia (fever, cough, dyspnea, rapid breathing but with regular arterial O_2_ saturation values); 3) Severe: patients with established lung damage, symptoms of pneumonia, cough, fever, dyspnea and hypoxia; and 4) Critical: patients with systemic extrapulmonary inflammation and the presence of large amounts of proinflammatory cytokines (World Health Organization, 2020a). Based on the fact that in most American countries, health policies restrict the diagnosis of COVID-19 to suspects with clear symptoms of infection and/or a proven history of contact with infected people (Aguirre, 2020; Americas Society Council of Administration, 2020; Galindo, 2020), in this study, we assumed that the cases observed and reported officially include mainly symptomatic (mild + moderate + severe + critical). We performed a theoretical calculation to determine the number of undiagnosed cases (asymptomatic + undiagnosed symptomatic). To do so, we used the infection mortality rates (IFR) of New York (IFR_NY_ = 1.4 - considered so far to be the most reliable calculated value available for this parameter (Worldometer, 2020a)) to calculate the “Estimated Total cases” (= observed deaths*IFR_NY_) (Jombart et al., 2020) and the “Estimated Fraction of Undiagnosed cases” (= [calculated total cases - observed cases]/calculated total cases) (Worldometer, 2020a); Verity et al. (2020)). Data on the observed deaths and recovered patients were obtained from each country’s official government website on May 23^rd^.

### 2.6. Evaluation of the impact of quarantines and self-isolation on the highland populations of Argentina, Bolivia, Colombia, Ecuador, and Peru

The impact of quarantines and self-isolation procedures was estimated for Argentina, Bolivia, Colombia, Ecuador, and Peru, as these countries applied early strict quarantines and self-isolation procedures. To do so, the number of people infected in a quarantine-free environment was estimated with SEIR epidemiological models (described above), using a “frequency of interaction” value equal to 30 (according to (Birbuet & López, 2020)). Next, these values were compared to the number of people infected obtained by using “frequency of interaction” values for each country calculated according to (Birbuet & López, 2020): Argentina (8.18), Bolivia (8.1), Colombia (9.03), Ecuador (8.7) and Peru (11.3).

## 3. Results

### 3.1. The incidence of COVID-19 is dependent on the population density in the American continent

The correlation between the number of positive cases and the population density for each location was evaluated at the continental level (23 countries). Our results show a significant positive correlation (Pearson’s correlation, *p* <0.0001; R2 = 0.004) between these two variables (S2). These results are in line with previous studies showing that the contagion rate and, therefore, the number of COVID-19 cases are positively correlated with population density. (Engle, Stromme, & Zhou, 2020; Jason Barr, 2020; Rocklöv & Sjödin, 2020; Rogers, 2020). As such, the number of COVID-19 cases normalized by population density (number of cases/population density) was used in the following analyses.

### 3.2. The incidence of COVID-19 decreases above 1,000 masl in the American countries

The correlation between the number of COVID-19 cases (normalized, logarithmized, and organized by intervals of 100 meters of altitude) and the altitude at the continental level (data from 23 countries together) was analyzed using a Pearson correlation analysis. Our results revealed a strong negative correlation (*p*<0.0001; R^2^= 0.603) between these variables, underlining a decrease in the incidence of COVID-19 cases with increasing altitude (see Fig. 1a). Furthermore, we remarked that a significant decrease in the number of normalized and logarithmized (but not yet organized by intervals of 100 meters of altitude) COVID-19 cases occurs already approximately 1,000 masl, with a further significant decrease at higher altitudes (Fig. 1b). Repeated correlation analysis performed at altitudes above 800, 1,000, 1,500 and 2,500 masl confirmed this observation. Nonsignificant correlation was found for data below 1,000 masl (Pearson correlation, *p*=0.568; R^2^= 0.042) (Fig. 1c), while a strongly significant correlation between COVID-19 incidence (ln [cases/population density]) and altitude was obtained above 1,000 masl (Pearson correlation, *p*<0.0001; R^2^= 0.427) (Fig. 1d). Furthermore, considering that various factors, such as public health policies, diagnostic strategies, confinement rules, and cultural aspects, may influence the number of reported cases of COVID-19, we performed a randomized block design ANOVA test to compare the number of COVID-19 cases (normalized, logarithmized, and organized by intervals of 100 meters of altitude) above and below 1,000 m. The advantage of this type of statistical analysis is that it considers the internal variability of each country in the incidence analysis. In line with our previous results, this test revealed a significant difference between the number of COVID-19 cases observed above versus below 1,000 masl (RBD-ANOVA F = 5,273; df= 45; *p* = 0.022;).

**Figure 1.**
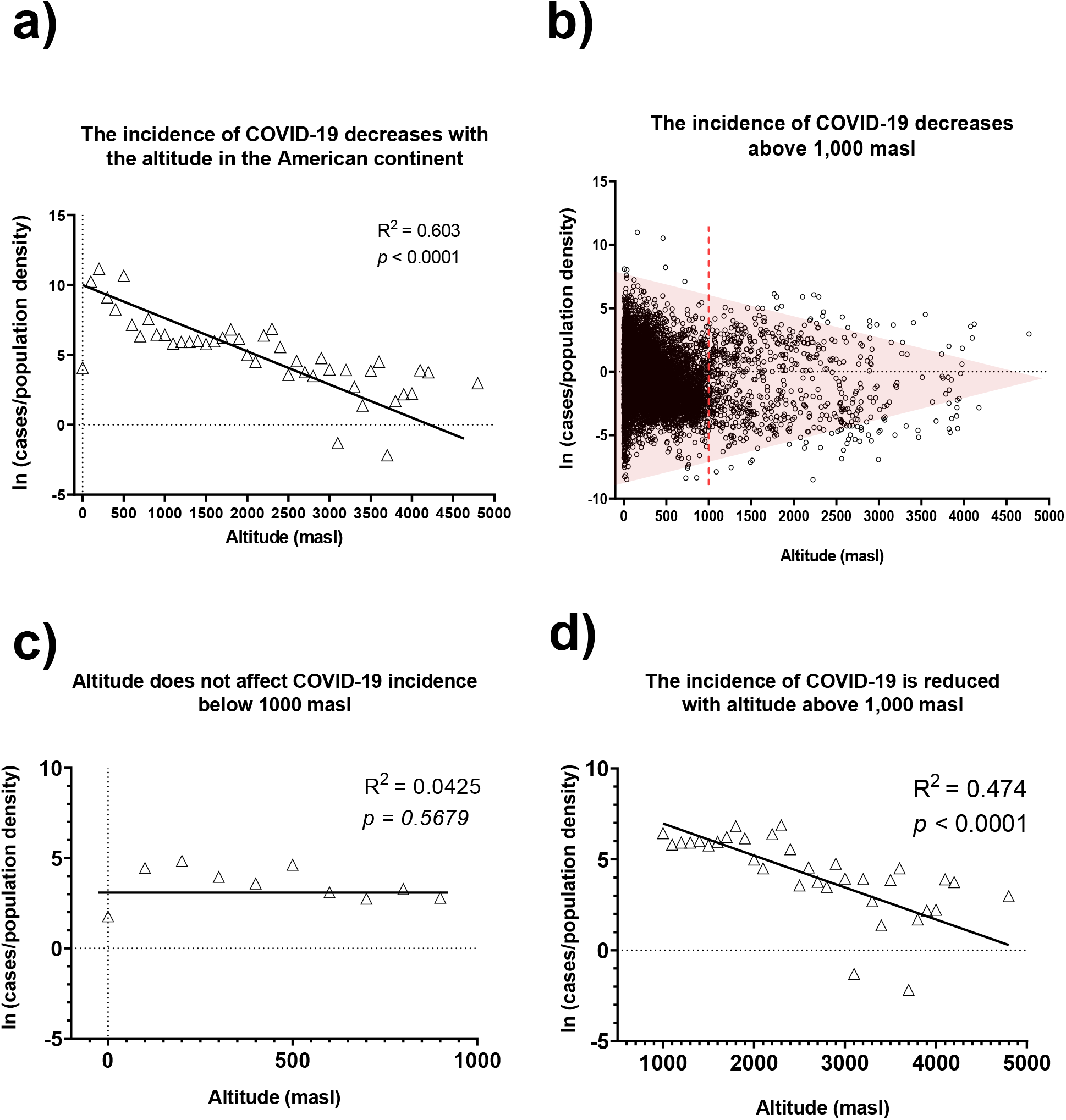
Correlation between population density and number of cases in the American continent. Epidemiological data were retrieved on May 23. Data on population density were extracted from the dataset created by CIESIN (Center for International Earth Science Information Network - CIESIN - Columbia University, 2018) or the corresponding country’s national statistics institute on May 23. Data were normalized by the population density of the same location and summed in intervals of 100 m of elevation. Raw, normalized, and adjusted data are available at https://doi.org/10.6084/m9.figshare.12685478 A) Correlation between altitude and the normalized (by population density) and adjusted (natural logarithm) number of positive COVID-19 cases in America grouped in intervals of 100 meters. B) Altitudinal distribution of the normalized and adjusted number of COVID-19 positive cases in the American continent. C) Correlation between altitude and the normalized and adjusted number of positive COVID-19 cases reported below 1,000 m in the American continent. D) Correlation between altitude and the normalized and adjusted number of positive COVID-19 cases reported above 1,0000 m in the American continent.

In the next step, we wanted to test whether the negative correlation found between the incidence of COVID-19 and the altitude for the American continent can be independently reproduced in each of the 14 American countries (of 23 analyzed) that reported cases at more than 1,000 masl. Our results showed significant negative correlations in 9 of 14 countries (Table 2) (S3): Argentina (*p*=0.003; R^2^= 0.597), Brazil (*p*<0.0001; R^2^= 0.789), Canada (*p*<0.012; R^2^= 0.012), Colombia (*p*<0.0001; R^2^= 0.376), Costa Rica (*p*=0.004; R^2^= 0.381), Ecuador (*p*<0.0001; R^2^= 0.647), Mexico (*p*=0.002; R^2^= 0.286), Peru (*p*=0.027; R^2^= 0.285) and USA (*p*<0.0001; R^2^= 0.577).

**Table 1.** Capitals of American countries located above 1,000 m above sea level

| City | Country | Population | Population density<br>(inhabitants/Km<sup>2</sup>) | Average altitude<br>(masl) |
| --- | --- | --- | --- | --- |
| La Paz | Bolivia | 1,864,191 | 378 | 3,600 |
| Quito | Ecuador | 1,100,000 | 475.51 | 2,850 |
| Bogota | Colombia | 6,422,198 | 24,643 | 2,625 |
| Mexico City | Mexico | 22,000,000 | 5,966 | 2,240 |
| Guatemala City | Guatemala | 1,700,000 | 159 | 1,529 |
| San Jose | Costa Rica | 2,723,000 | 7,548 | 1,146 |
| Brasilia | Brazil | 1,650,000 | 423.28 | 1,079 |

**Table 2.** Correlation between altitude and the incidence of COVID-19 in American countries.

| Country | Max. – Min. altitude<br>w/reported cases (masl) | R<sup>2</sup> | Pearson<br>correlation *p* |
| --- | --- | --- | --- |
| Argentina | 20 – 3,211 | 0.597 | **0.003** |
| Belize | 0 – 654 | 0.219 | 0.690 |
| Bolivia | 109 – 4,176 | 0.036 | 0.343 |
| Brazil | 0 – 1,615 | 0.789 | **<0.0001** |
| Canada | 1 – 2,115 | 0.068 | **0.012** |
| Chile | 351 – 3,169 | 0.058 | 0.405 |
| Colombia | 0 – 3,629 | 0.376 | **<0.0001** |
| Costa Rica | 0 – 2,077.78 | 0.381 | **0.004** |
| Cuba | 18 - 315.77 | 0.219 | 0.673 |
| Dominican Republic | 15 – 1,113 | 0.14 | 0.321 |
| Ecuador | 4 – 3,976 | 0.647 | **<0.0001** |
| El Salvador | 3 – 788 | 0.005 | 0.862 |
| French Guyana | 187 – 187 | Not enough data | Not enough data |
| Haiti | 0 – 1,499 | 0.313 | 0.149 |
| Honduras | 4 – 1,689 | 0.079 | 0.374 |
| Mexico | 1 – 2,593 | 0.286 | **0.002** |
| Panama | 0 – 1,402 | 0.154 | 0.184 |
| Paraguay | 61 – 501 | 0.630 | 0.109 |
| Peru | 7 – 4,765 | 0.285 | **0.027** |
| Puerto Rico | 0 – 846 | 0.15 | 0.3432 |
| Uruguay | 27 – 163 | Not enough data | Not enough data |
| USA | 0 – 3,510 | 0.577 | **<0.0001** |
| Venezuela | 9 – 1,907 | 0.165 | 0.1907 |

Taken together, these data confirm our previous observations (Arias-Reyes et al., 2020), suggesting that the virulence of SARS-CoV-2 gradually decreases with altitude. However, although we previously suggested that the incidence of COVID-19 decreases at high altitude (2,500 masl), the data of the present study show a significant decrease in the disease incidence starting at 1,000 m of altitude.

### 3.3. The transmission rate of SARS-CoV-2 is lower in the highlands compared to lowlands

To investigate whether the transmission rate of SARS-CoV-2 differs between highlands (>1,000 masl) and lowlands (<1,000 masl), we used SEIR epidemiological models only in countries that applied similar strong early quarantines and provided daily epidemiological data at state/province/departamento level: Argentina, Bolivia, Colombia, Ecuador, and Peru. To do so, the “transmission rate” values that allowed the most faithful theoretical reproduction of the epidemiological curves observed in the highlands and lowlands were obtained (Fig. 2). Our results show that compared to lowlands (Argentina=4.2%, Bolivia=4%, Colombia 3.7%, Ecuador=3.8%, Peru=4%), lower “transmission rates‥ at highlands lead to better modeling data of the real epidemiological curves (Argentina=2.5%, Bolivia=3%, Colombia 3.6%, Ecuador=3.6%, Peru=3%). These results strongly support the hypothesis of decreased SARS-CoV-2 virulence in highlands compared to lowlands.

**Figure 2.**
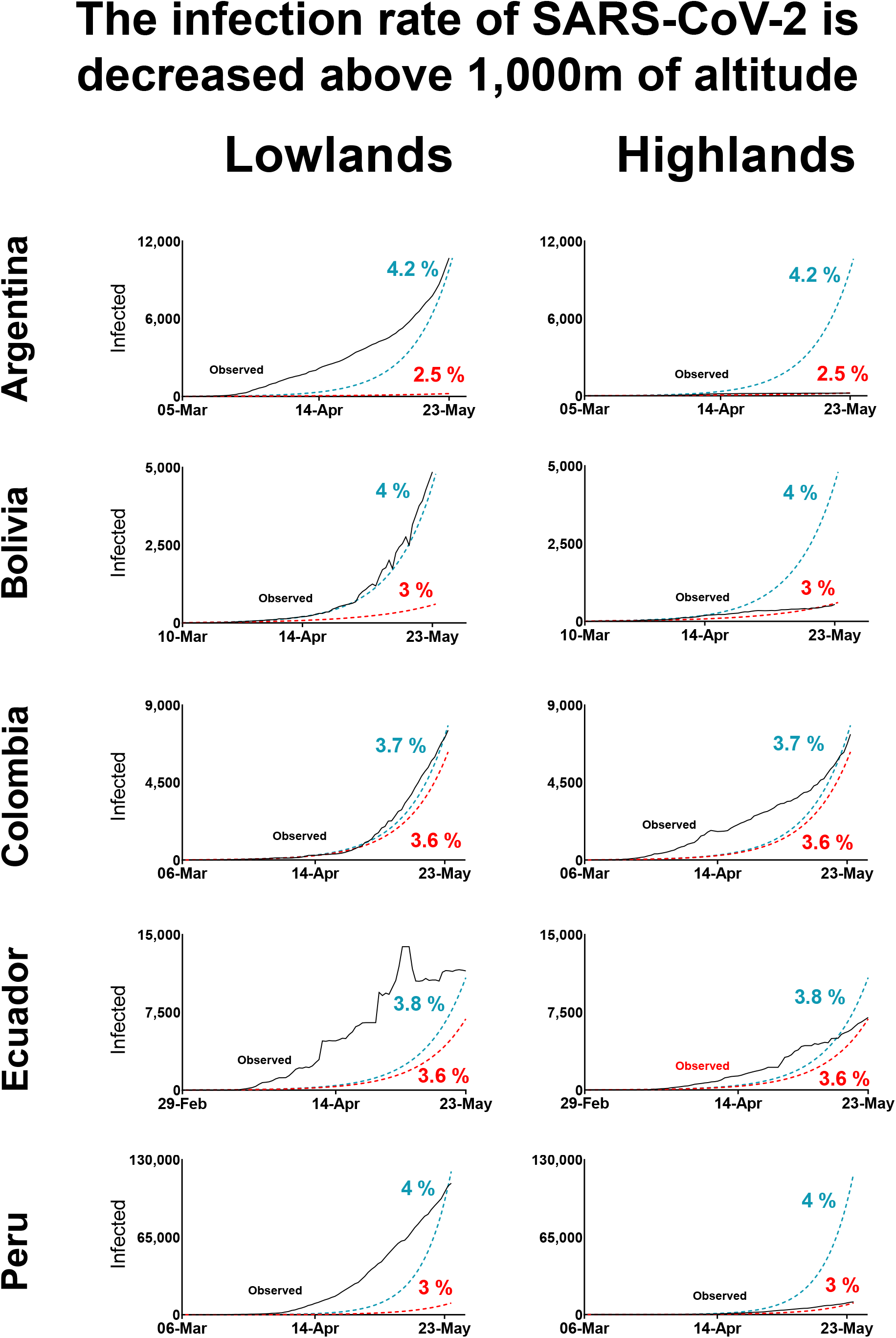
Effect of the transmission rate in the epidemiological pattern of COVID-19 in lowlands and highlands in Argentina, Bolivia, Colombia, Ecuador, and Peru, where strict early quarantines were applied and daily epidemiological data at state/province/departamento were available by May 23. For each country, the black lines show the observed cases in lowlands or highlands. The red dotted lines represent the modeled data using the best-fit value of “transmission rate” calculated for highlands. The blue dotted lines represent the modeled data using the best-fit value of “transmission rate” for lowlands. The red dotted lines represent the modeled data using the best-fit value of “transmission rate” calculated for highlands.

**Figure 3.**
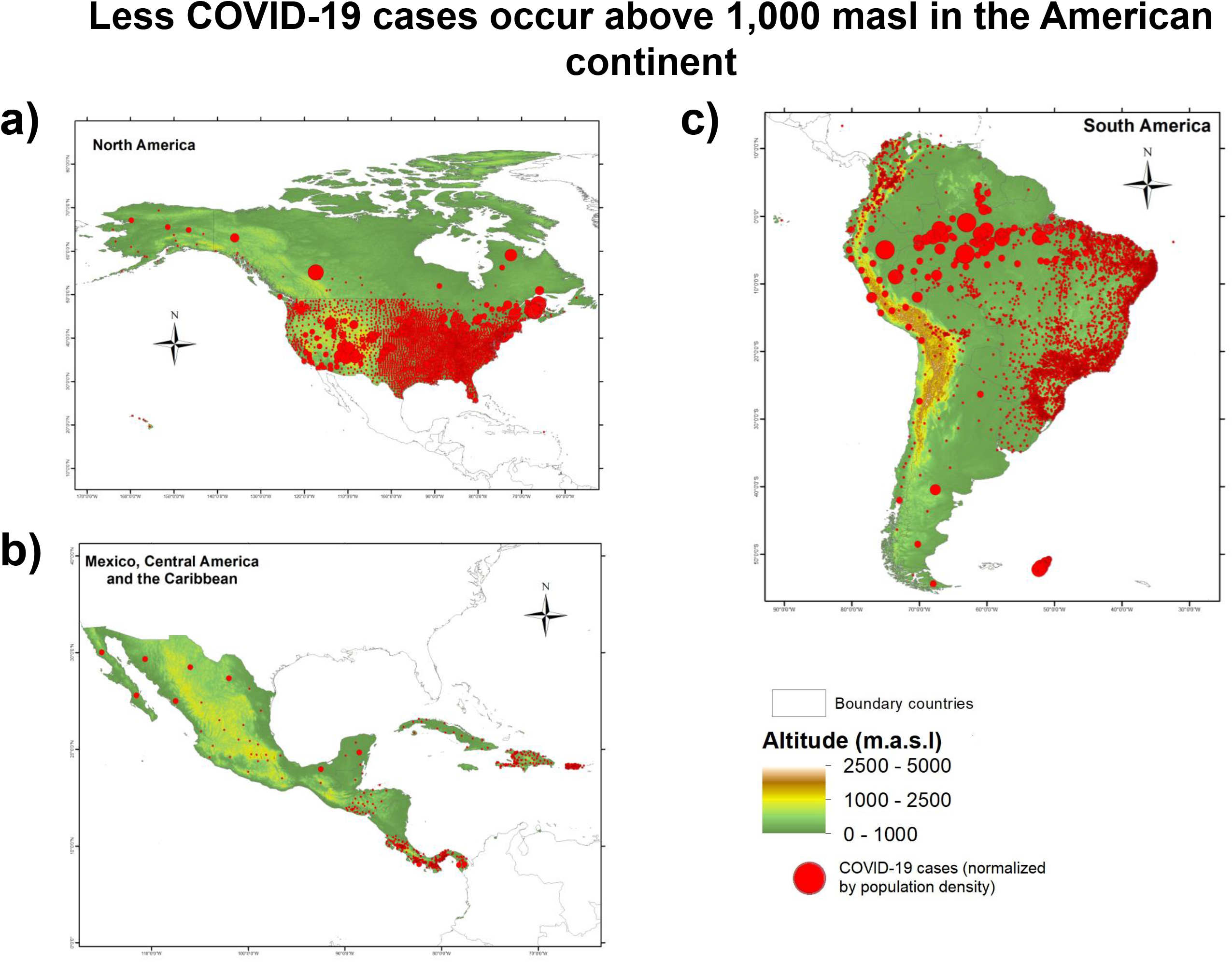
Geographic and altitudinal distribution COVID-19 in A) North America, B) Central America and C) South America. Blue circles represent COVID-19 positive cases; the radius of the circle is relative to the normalized number of cases (cases/population density) in the location. The geographical coordinates and epidemiological data were retrieved on May 23 as described in the methods section. The final database is available at https://doi.org/10.6084/m9.figshare.12685478 Maps for the 23 studied countries are available at https://doi.org/10.6084/m9.figshare.12685664.v1

Of note, the “transmission rate” values between the highlands and the lowlands in Colombia show a small difference. This result is explained by the fact that 60% of its population (∼ 30 million inhabitants) is settled above 1,000 m altitude, which are also the most densely populated areas. This causes the requirement of a higher infection probability value in the SEIR model to fit the real data, thus approaching the values of the transmission rate between highlands and lowlands.

### 3.4 The severity of COVID-19 is reduced in highlands compared to lowlands

Classically, the estimation of the severity of a disease is performed by a comparison between recovery rates and infection mortality rates (IFR). However, since this information is still limited for COVID-19, we used a theoretical rationalization approach to estimate this parameter. We evaluated the differences in the percentage of recovered patients (from the total reported cases) and in the death-to-case ratio (deaths/total reported cases) as indicators of the recovery rate and the IFR for Argentina, Bolivia, Colombia, Ecuador, and Perú (countries that applied similar strong early quarantines and provided daily epidemiological data at the state/province/departamento level). We found a significantly higher percentage of recovered patients in the highlands (Wilcoxon signed-rank test *p*= 0.031) versus lowlands, suggesting higher a recovery rate in the highlands versus lowlands. On the other hand, our results did not show significant differences of death-to-case ratio between the highlands and the lowlands (Table 3). This may occur only in the case in which the number of undiagnosed cases (asymptomatic + undiagnosed symptomatic) is not similar between the highlands and lowlands. Indeed, our theoretical approach for the assessment of undiagnosed cases showed approximately 76% of undiagnosed cases in the highlands and 73% of undiagnosed cases in the lowlands (1.04 ± 0.12-fold higher) (Table 4).

**Table 3.** Percentage of recovered and death rates in COVID-19 patients.

| Country | Percentage of Recovered Patients |  | Death-to-case-ratio |  |
| --- | --- | --- | --- | --- |
|  | Highlands (%) | Lowlands (%) | Highlands (%) | Lowlands (%) |
| Argentina | 57,57 | 36,05 | 4,66 | 4,95 |
| Bolivia | 35,73 | 5,62 | 5,79 | 3,84 |
| Colombia | 4,05 | 3,46 | 36,42 | 15,7 |
| Ecuador | 9,1 | 2,38 | 9,56 | 36,0 |
| Peru | 25,77 | 8,22 | 3,5 | 3,70 |

**Table 4.** Estimated percentage of undiagnosed COVID-19 cases in five American countries.

| Country | Highlands (%) | Lowlands (%) |
| --- | --- | --- |
| Argentina | 69.2 | 64.4 |
| Bolivia | 74.8 | 61.9 |
| Colombia | 95.5 | 88.5 |
| Ecuador | 85.5 | 96.6 |
| Peru | 58.4 | 56.0 |
| **Mean** | 75.8 | 73.5 |
| **S.D.** | 15.9 | 17.9 |

Although, these values are theoretical (and should be interpreted carefully), they suggest that the severity of COVID-19 is lower at highlands compared to lowlands.

### 3.5 Strict quarantine and self-isolation measures at altitude significantly decreased the rapid progression of COVID-19

To close our analysis, we wanted to assess the impact of quarantine and social distancing measures in countries that applied early strict measurements, such as Argentina, Bolivia, Colombia, Ecuador, and Peru. By using SEIR epidemiological models, we compared the number of people infected under conditions in which the “frequency of interaction” value is associated with a quarantine-free environment (= 30) to the number of infected people under conditions in which the “frequency of interaction” value is associated with a strict quarantine environment: 8.18 for Argentina, 8.1 for Bolivia, 9.03 for Colombia, 8.7 for Ecuador, and 11.3 for Peru (11.3). Our results show that the application of containment measures drastically delayed the number of COVID-19 infections (Figure 4).

**Figure 4.**
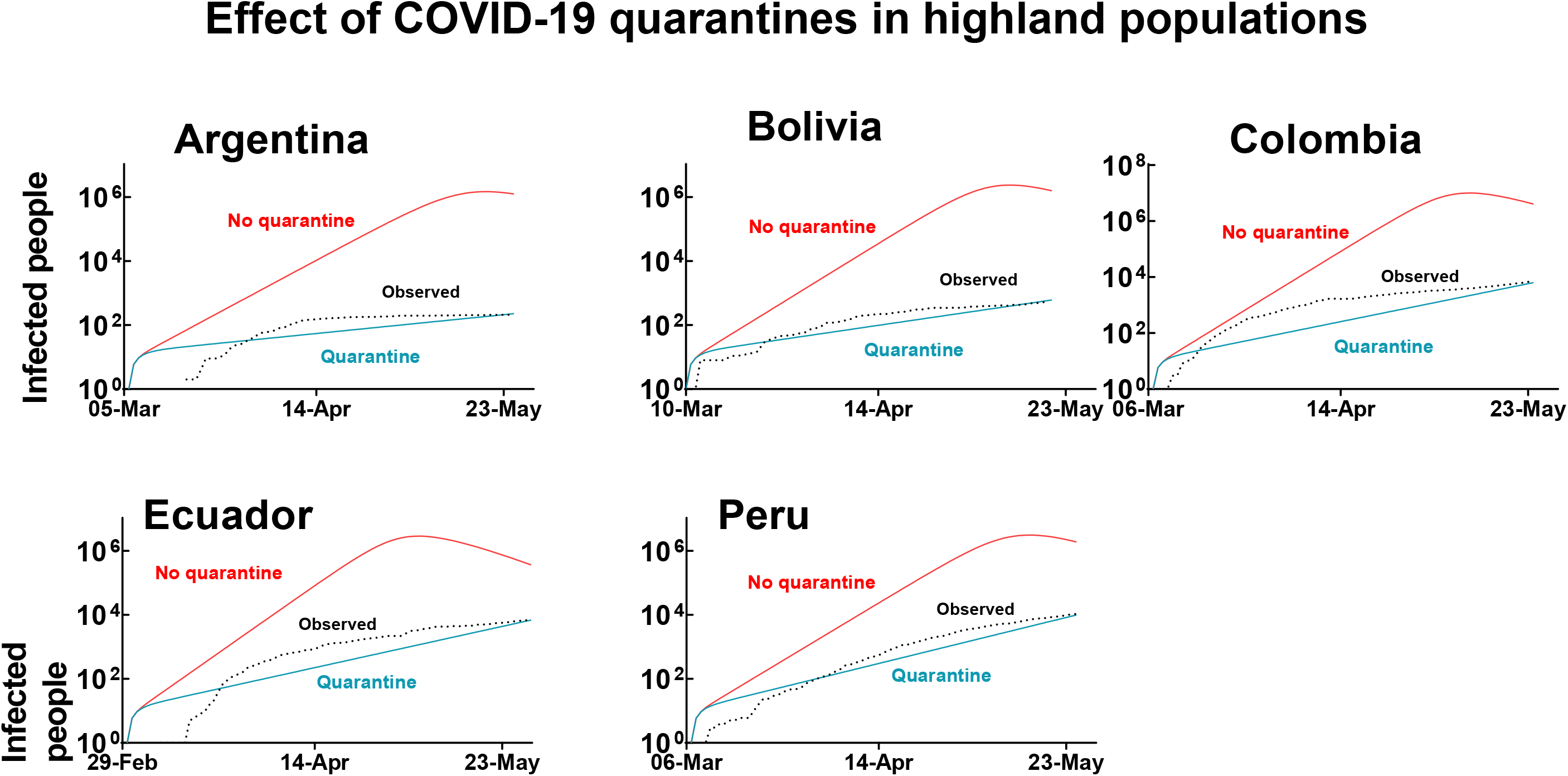
Effect of quarantines in highlands populations of Argentina, Bolivia, Colombia, Ecuador, and Peru, where strict early quarantines were applied. For each country, the black dotted lines show the observed numbers of infected people since the beginning of the pandemic. The blue lines show the numbers of infected people calculated with the SEIR model considering the application of quarantines. The red lines show the numbers of infected people estimated with the SEIR model considering quarantines would not be applied.

## 4. Discussion

By expanding our analysis to 23 countries of the American continent, this work clearly demonstrates our previous observations suggesting that the virulence of SARS-CoV-2 decreases significantly with altitude. Indeed, in a previous study we reported that global data analysis (that included detailed information from the Tibetan Autonomous Region of China, Bolivia, and Ecuador) suggested that COVID-19 decreases significantly at high altitude (2,500 masl) (Arias-Reyes, et al 2020). While subsequent reports from other teams confirmed this observation (Accinelli & Leon-Abarca, 2020; Huamaní et al., 2020; Ortiz-Prado et al., 2020; Rivero & Montoya, 2020), in this new study we appropriately corrected the COVID-19 data by population density. Remarkably, our results show that a clear turning point in the incidence of the disease occurs at 1,000 masl. Moreover, independent examination from all the American countries that reported cases at more than 1,000 masl (14 out of 23) confirmed this result, except for four nations: Bolivia, in which the low number of cases reported for moderate-altitude regions (1,000 – 2,500 masl) masked a possible correlation; Chile, in which its long and narrow geography (flanked by the Andean mountains on the east and the Pacific Ocean) does not allow obtaining precise altitude values for COVID-19 cases; and Dominican Republic, Haiti and Panamá, in which the highest altitude with reported cases is just over 1,000 masl. Therefore, these data may have a low weight in the statistical evaluation.

To better understand this result, we performed additional theoretical approaches to determine the capability of the virus to move from one host to another (the transmission rate of the virus). Our results show that this parameter decreases significantly in the highlands compared to lowlands, suggesting that environmental factors influence the virulence of SARS-CoV-2 at above 1000 m. Indeed, as altitude increases, the environment is characterized by more drastic changes in temperature between night and day, higher air dryness, and higher levels of ultraviolet (UV) light radiation. (United-States-Environmental-Protection-Agency, 2017). In particular, UV light radiation was suggested to be an important natural sanitizer at altitude that may shorten the half-life of any given virus (Andrade, 2020; G. Zubieta-Calleja, 2020; G. R. Zubieta-Calleja & Zubieta-DeUrioste, 2017). In addition, the solar radiation is also more intense at altitude, and a recent study reported that this factor may also be a key factor leading to the deactivation of the virus (Asyary & Veruswati, 2020; Li, Li, Zhang, & Liu, 2020). Taken together, these factors may lead to a gradual reduction of the “survival” and “virulence” capacity of the virus as altitude progresses. Finally, the size of the virus inoculum in the air should gradually decrease as the barometric pressure decreases and the distance between the air molecules increases.

Apart from environmental factors that may decrease the transmission capacity of the virus, our results also suggest that physiological mechanisms associated with a prolonged exposure to barometrical hypoxia help to decrease the severity of the infection. Thus far, two factors have been suggested that may be involved in this phenomenon: A decreased expression of the virus’s gateway to the body, angiotensin converting enzyme 2 (ACE2) (Arias-Reyes, et al, 2020), and an increased level of erythropoietin (EPO) (Soliz, et al., 2020). Stabilization of the hypoxia inducible factor 1 alpha (HIF1-a - a master regulator of the response to hypoxia) may lead the regulation of both parameters. Indeed, it was shown that exposure of human pulmonary artery smooth muscle cells (hPASMC) to hypoxia markedly decreases the expression of ACE2 (Zhang et al., 2009). Similar results were obtained in heart cells of Sprague Dawley rats after 28 days exposure to conditions equivalent to 10% O2 hypoxia (Dang et al., 2020). Furthermore, it was shown that changes in oxygen availability as small as 2% are sufficient to induce HIF activation in many tissues (Jiang, Semenza, Bauer, & Marti, 1996; Stroka et al., 2001). In the same direction, HIF is the main booster of EPO production in the kidney and other tissues, and it was observed that the HIF-related oxygen sensing mechanism precisely senses altitude differences of 300 m even in low to moderate altitudes (Gassmann et al., 2019). While EPO is the central factor leading the stimulation of red blood cells (Martin & Windsor, 2008; Storz & Moriyama, 2008), EPO also promotes adaptive cellular responses to hypoxic challenges and tissue-damaging insults in nonhematopoietic tissues (Juul & Felderhoff-Mueser, 2007). As such, in the infectious context induced by the SARS-CoV-2 virus, since EPO may help improve oxygen delivery to the tissues, it may also protect other tissues from a multiple organ dysfunction and inflammation (Chen et al., 2020; Yang et al., 2020).

Although the results of this work clearly indicate that the virulence of SARS-CoV-2 decreases with altitude, these results should not be misinterpreted either by citizens or by government systems responsible for imposing sanitary restrictions. Indeed, to make this point clear, we performed calculations to determine the impact of quarantine and self-isolation measures in highlands of countries that applied early strict measurements, such as Argentina, Bolivia, Colombia, Ecuador, and Peru. The results of this analysis show that without relevant restriction measures, the number of currently reported cases of COVID-19 would be increased by 10,000 times (Figure 4).

Finally, the limitations of the study include possible failures and delays in reporting cases as well as that our analysis does not consider the effect of different risk groups such as age, sex, comorbidities, and the possibility of reinfection.

In conclusion, epidemiological analyses of the present work strongly suggest that the incidence of COVID-19 significantly decreases with altitude with a turning point at 1,000 masl. This effect seems to be related both to a decrease in the transmission capacity of the virus and to the physiological characteristics of altitude residents. Finally, our results suggest that knowledge of the mechanisms of physiological acclimatization to hypoxia may help to better understand the viral nature of SARS-CoV-2 as well as facilitate the discovery of new treatment strategies.

## Data Availability

Raw, normalized, and adjusted data of COVID-19 cases are available and registered at figshare.
Raw epidemiological daily data of COVID-19 of Argentina, Bolivia, Colombia, Ecuador, and Peru are available and registered at figshare.

https://doi.org/10.6084/m9.figshare.12685523

https://doi.org/10.6084/m9.figshare.12685478

## DATA AVAILABILITY

Raw, normalized, and adjusted data of COVID-19 cases are available at https://doi.org/10.6084/m9.figshare.12685478

Raw epidemiological daily data of COVID-19 of Argentina, Bolivia, Colombia, Ecuador, and Peru are available at https://doi.org/10.6084/m9.figshare.12685523

## AUTHOR CONTRIBUTIONS

C.A.-R., L.P.-M., F.A.-R., and F.C.-R. gathered the epidemiological and geographical data. F.C.- R. processed the geographical data and created the distribution maps. C.A.-R. and J.S. analyzed the data. J.S., C.A.-R., L.P.-M., F.A.-R, D.A.M., N. Z.D., R.A.-T., E.M.S.-G., G.Z.-C., and M.D. wrote this manuscript.

## ACKNOWLEDGMENTS

The authors wish to acknowledge Dr Gonzalo Leonardini for his advice and assistance in the mathematical evaluation of our models, the platform CovidBot Peru for giving us access to their database of systematized daily epidemiological information from Peru, and Teresa Roncal for her inputs and thoughts on the redaction of the manuscript.

## Supplementary material

**S1**. Table of sources of epidemiological data

**S2**. Effect of population density on the incidence of COVID-19 cases in the American continent. Epidemiological data were retrieved on May 23. Data on population density were extracted from the dataset created by CIESIN (Center for International Earth Science Information Network - CIESIN - Columbia University, 2018) or the corresponding country’s national statistics institute on May 23.

**S3**. Effect of altitude on the incidence of COVID-19 cases in American countries. Epidemiological data were retrieved on May 23. Data on population density were extracted from the dataset created by CIESIN (Center for International Earth Science Information Network - CIESIN - Columbia University, 2018) or the corresponding country’s national statistics institute on May 23. Data were normalized by the population density of the same location, adjusted by calculating the natural logarithm (ln) of each value, and summed in intervals of 100 m of elevation. Raw, normalized, and adjusted data are available at https://doi.org/10.6084/m9.figshare.12685478

## REFERENCES

Abadie, A., Bertolotti, P., & Arnab, B. D. (2020). Epidemic Modeling and Estimation.

Accinelli, R. A., & Leon-Abarca, J. A. (2020). En la altura el COVID-19 es menos frecuente: la experiencia del Perú. Archivos de Bronconeumología.

Aguirre, M. (2020). Las dudas sobre el número de pruebas de Covid-19 en América Latina. Retrieved from https://www.france24.com/es/20200509-dudas-numero-pruebas-covid-19-america-latina

Alcaldía de Bogotá. (2020). Cifras de coronavirus (COVID-19) en Bogotá. Retrieved from https://bogota.gov.co/mi-ciudad/salud/coronavirus/cifras-de-coronavirus-covid-19-en-bogota

Americas Society Council of Administration, A. (2020). Where Is the Coronavirus in Latin America? |AS/COA. Retrieved from https://www.as-coa.org/articles/where-coronavirus-latin-america

Andrade, M. (2020). La radiación ultravioleta en tiempos de cuarentena. Pagina siete, Bolivia, https://www.paginasiete.bo/opinion/2020/4/10/la-radiacion-ultravioleta-en-tiempos-de-cuarentena-252240.html.

Arias-Reyes, C., Zubieta-DeUrioste, N., Poma-Machicao, L., Aliaga-Raduan, F., Carvajal-Rodriguez, F., Dutschmann, M., … Soliz, J. (2020). Does the pathogenesis of SARS-CoV-2 virus decrease at high-altitude? Respiratory physiology & neurobiology, 277, 103443. doi:https://doi.org/10.1016/j.resp.2020.103443

Asyary, A., & Veruswati, M. (2020). Sunlight exposure increased Covid-19 recovery rates: A study in the central pandemic area of Indonesia. Science of The Total Environment, 729, 139016. doi:https://doi.org/10.1016/j.scitotenv.2020.139016

Birbuet, J. C., & López, R. (2020). Dinámica de expansión del COVID-19 en Bolivia durante las primeras 6 semanas.

Center for International Earth Science Information Network - CIESIN - Columbia University. (2018). Gridded Population of the World, Version 4 (GPWv4): Population Density, Revision 11. Retrieved from: https://doi.org/10.7927/H49C6VHW

Chen, N., Zhou, M., Dong, X., Qu, J., Gong, F., Han, Y., Zhang, L. (2020). Epidemiological and clinical characteristics of 99 cases of 2019 novel coronavirus pneumonia in Wuhan, China: a descriptive study. Lancet, 395(10223), 507–513. doi:10.1016/S0140-6736(20)30211-7

Dang, Z., Su, S., Jin, G., Nan, X., Ma, L., Li, Z., Ge, R. (2020). Tsantan Sumtang attenuated chronic hypoxia-induced right ventricular structure remodeling and fibrosis by equilibrating local ACE- AngII-AT1R/ACE2-Ang1-7-Mas axis in rat. J Ethnopharmacol, 250, 112470. doi:10.1016/j.jep.2019.112470

Engle, S., Stromme, J., & Zhou, A. (2020). Staying at home: mobility effects of covid-19. Available at SSRN.

Fick, S. E., & Hijmans, R. J. (2017). WorldClim 2: new 1-km spatial resolution climate surfaces for global land areas. International Journal of Climatology, 37(12), 4302–4315. doi:10.1002/joc.5086

France 24. (2020, 2020-05-04). France sticks by lockdown easing plan despite mayors’ ‘forced march’ concerns. Retrieved from https://www.france24.com/en/20200504-france-sticks-by-lockdown-easing-plan-despite-mayors-forced-march-concerns

Galindo, J. (2020). Faltan pruebas para medir el virus (y muchos casos por contar) en Latinoamérica. Retrieved from https://elpais.com/sociedad/2020-04-20/faltan-pruebas-para-medir-el-virus-y-muchos-casos-por-contar-en-latinoamerica.html

GmbH, O. (2020). OpenCage Geocoder. Retrieved from https://opencagedata.com/

Gobierno de la Ciudad de México. (2020). Datos abiertos sobre salud pública, acciones sociales y gasto público en la Ciudad de México. Retrieved from https://datos.cdmx.gob.mx/pages/covid19/

Hornbein, T., & Schoene, R. (2001). High altitude: an exploration of human adaptation: CRC Press.

Huamaní, C., Velásquez, L., Montes, S., & Miranda-Solis, F. (2020). Propagation by COVID-19 at high altitude: Cusco case. Respiratory physiology & neurobiology, 279, 103448–103448. doi:10.1016/j.resp.2020.103448

Jason Barr, T. T. (2020). Are Crowded Cities the Reason for the COVID-19 Pandemic? Retrieved from https://blogs.scientificamerican.com/observations/are-crowded-cities-the-reason-for-the-covid-19-pandemic/

Jiang, B.-H., Semenza, G. L., Bauer, C., & Marti, H. H. (1996). Hypoxia-inducible factor 1 levels vary exponentially over a physiologically relevant range of O2 tension. American Journal of Physiology-Cell Physiology, 271(4), C1172–C1180.

Jombart, T., van Zandvoort, K., Russell, T., Jarvis, C., Gimma, A., Abbott, S., … Edmunds, W. (2020). Inferring the number of COVID-19 cases from recently reported deaths [version 1; peer review: 2 approved]. Wellcome Open Research, 5(78). doi:10.12688/wellcomeopenres.15786.1

Joshua Berlinger, C. N. N. (2020). Coronavirus has now spread to every continent except Antarctica. Retrieved from https://www.cnn.com/2020/02/25/asia/novel-coronavirus-covid-update-us-soldier-intl-hnk/index.html

Juul, S., & Felderhoff-Mueser, U. (2007). Epo and other hematopoietic factors. Semin Fetal Neonatal Med, 12(4), 250–258. doi:10.1016/j.siny.2007.01.015

Langton, K. (2020, 2020-05-06). China lockdown: How long was China on lockdown? Express. Retrieved from https://www.express.co.uk/travel/articles/1257717/china-lockdown-how-long-was-china-lockdown-timeframe-wuhan

Li, Y., Li, Q., Zhang, N., & Liu, Z. (2020). Sunlight and vitamin D in the prevention of coronavirus disease (COVID-19) infection and mortality in the United States.

Martin, D., & Windsor, J. (2008). From mountain to bedside: understanding the clinical relevance of human acclimatisation to high-altitude hypoxia. Postgrad Med J, 84(998), 622-627; quiz 626. doi:10.1136/pgmj.2008.068296

Ministerio de Saude de Brasil. (2020). Coronavirus Brasil. Retrieved from https://covid.saude.gov.br/

Montréal, S. (2020). CORONAVIRUS COVID-19 - Current situation in Montréal. Retrieved from https://santemontreal.qc.ca/en/public/coronavirus-covid-19/

N. Y. C. Health Department. (2020). COVID-19: Data Summary - NYC Health. Retrieved from https://www1.nyc.gov/site/doh/covid/covid-19-data.page

NPR. (2020). Millions Return To Work In Italy After Weeks Of Lockdown. Retrieved from https://www.npr.org/sections/coronavirus-live-updates/2020/05/04/849989973/millions-return-to-work-in-italy-after-weeks-of-lockdown

Ortiz-Prado, E., Diaz, A. M., Barreto, A., Moyano, C., Arcos, V., Vasconez-Gonzalez, E., … Fernandez-Naranjo, R. (2020). Epidemiological, socio-demographic and clinical features of the early phase of the COVID-19 epidemic in Ecuador. medRxiv.

Pan-American Health Organization. (2020). Coronavirus Disease (COVID-19) - PAHO/WHO |Pan American Health Organization. Retrieved from https://www.paho.org/en/topics/coronavirus-infections/coronavirus-disease-covid-19

Rede Brasil Atual. (2020). Casos de covid-19 na periferia de São Paulo disparam 45% em uma semana. Retrieved from https://www.redebrasilatual.com.br/saude-e-ciencia/2020/04/casos-covid-19-periferia-sp/

Rivero, A. C., & Montoya, M. (2020). COVID19 EN POBLACIÓN RESIDENTE DE ZONAS GEOGRÁFICAS A ALTURAS SUPERIORES A 2500 msnm. SITUA, 23(1), 16–16.

Rocklöv, J., & Sjödin, H. (2020). High population densities catalyse the spread of COVID-19. J Travel Med, 27(3). doi:10.1093/jtm/taaa038

Rogers, A. (2020). How Does a Virus Spread in Cities? It’s a Problem of Scale. Wired. Retrieved from https://www.wired.com/story/how-does-a-virus-spread-in-cities-its-a-problem-of-scale

SEDES La Paz. (2020). Información Oficial SEDES La Paz situación COVID-19. Retrieved from https://www.sedeslapaz.gob.bo/reportes_covid19

Storz, J. F., & Moriyama, H. (2008). Mechanisms of hemoglobin adaptation to high altitude hypoxia. High Alt Med Biol, 9(2), 148–157. doi:10.1089/ham.2007.1079

Stroka, D. M., Burkhardt, T., Desbaillets, I., Wenger, R. H., Neil, D. A., Bauer, C., … Candinas, D. (2001). HIF-1 is expressed in normoxic tissue and displays an organ-specific regulation under systemic hypoxia. The FASEB Journal, 15(13), 2445–2453.

United-States-Environmental-Protection-Agency. (2017). Calculating the UV Index. https://www.epa.gov/sunsafety/calculating-uv-index-0.

Verity, R., Okell, L. C., Dorigatti, I., Winskill, P., Whittaker, C., Imai, N., … Ferguson, N. M. (2020). Estimates of the severity of coronavirus disease 2019: a model-based analysis. The Lancet Infectious Diseases, 20(6), 669–677. doi:10.1016/S1473-3099(20)30243-7

World Health Organization. (2020a). Clinical management of COVID-19: interim guidance, 27 May 2020. Retrieved from Geneva: https://apps.who.int/iris/handle/10665/332196

World Health Organization. (2020b). Coronavirus Disease (COVID-19) - events as they happen. Retrieved from https://www.who.int/emergencies/diseases/novel-coronavirus-2019/events-as-they-happen

Worldometer. (2020a). Coronavirus Death Rate (COVID-19) - Worldometer. Retrieved from https://www.worldometers.info/coronavirus/coronavirus-death-rate/

Worldometer. (2020b). Coronavirus Update (Live): 3,804,912 Cases and 263,349 Deaths from COVID-19 Virus Pandemic - Worldometer. Retrieved from https://www.worldometers.info/coronavirus/

Yang, X., Yu, Y., Xu, J., Shu, H., Xia, J., Liu, H., … Shang, Y. (2020). Clinical course and outcomes of critically ill patients with SARS-CoV-2 pneumonia in Wuhan, China: a single-centered, retrospective, observational study. Lancet Respir Med. doi:10.1016/S2213-2600(20)30079-5

Zhang, R., Wu, Y., Zhao, M., Liu, C., Zhou, L., Shen, S., … Wan, H. (2009). Role of HIF-1alpha in the regulation ACE and ACE2 expression in hypoxic human pulmonary artery smooth muscle cells. Am J Physiol Lung Cell Mol Physiol, 297(4), L631–640. doi:10.1152/ajplung.90415.2008

Zubieta-Calleja, G. (2020). Covid-19 Pandemia Essential Suggestions. https://altitudeclinic.com/blog/2020/03/covid-19-pandemia-essential-suggestions/,.

Zubieta-Calleja, G. R., & Zubieta-DeUrioste, N. A. (2017). Extended longevity at high altitude: Benefits of exposure to chronic hypoxia. BLDE University Journal of Health Sciences, 2(2), 80–90.

